# The Zero-Corrected, Gravity-Model Estimator (ZERO-G): A novel method to create high-quality, continuous incidence estimates at the community-scale from passive surveillance data

**DOI:** 10.1101/2023.03.13.23287196

**Authors:** Michelle V Evans, Felana A Ihantamalala, Mauricianot Randriamihaja, Andritiana Tsirinomen’ny Aina, Matthew H Bonds, Karen E Finnegan, Rado JL Rakotonanahary, Mbolatiana Raza-Fanomezanjanahary, Bénédicte Razafinjato, Oméga Raobela, Sahondraritera Herimamy Raholiarimanana, Tiana Harimisa Randrianavalona, Andres Garchitorena

## Abstract

Data on population health are vital to evidence-based decision making but are rarely adequately localized or updated in continuous time. They also suffer from low ascertainment rates, particularly in rural areas where barriers to healthcare can cause infrequent touch points with the health system. Here, we demonstrate a novel statistical method to estimate the incidence of endemic diseases at the community level from passive surveillance data collected at primary health centers. The zero-corrected, gravity-based (ZERO-G) estimator explicitly models sampling intensity as a function of health facility characteristics and statistically accounts for extremely low rates of ascertainment. The result is a standardized, real-time estimate of disease incidence at a spatial resolution nearly ten times finer than typically reported by facility-based passive surveillance systems. We assessed the robustness of this method by applying it to a case study of field-collected malaria incidence rates from a rural health district in southeastern Madagascar. The ZERO-G estimator decreased geographic and financial bias in the dataset by over 90% and doubled the agreement rate between spatial patterns in malaria incidence and incidence estimates derived from prevalence surveys. The ZERO-G estimator is a promising method for adjusting passive surveillance data of common, endemic diseases, increasing the availability of continuously updated, high quality surveillance datasets at the community scale.

## INTRODUCTION

Health metrics are vital to public health efforts, allowing decision makers to better understand the state of population health and evaluate the impact of health interventions ^1,2^. Many of these metrics are based on routine passive disease surveillance from facility-based health management information systems (HMIS), which record the number of disease cases received at each facility at a regular frequency. Health records are then aggregated, digitized, and transferred to the district and, eventually, national health offices ^3^. While the exact structure differs by country, the scale of spatial aggregation of the data in an HMIS corresponds to the specific level of the health system and its corresponding health infrastructure. For example, national-level data are used by international organizations to monitor long-term, multi-country trends and inform policy; regional- and district-level surveillance data may be used by national public health offices to allocate resources within the country; and individual health facility information is used by district health offices for program management.

Missing from most HMIS are routine surveillance data at the scale of individual communities or villages. These data are needed for spatially targeted interventions for disease control in collaboration with community health programs, which primarily serve rural communities and play an integral role in achieving universal health coverage ^4,5^. While rural primary care facilities typically serve over ten thousand people spread along hundreds of square kilometers, community health workers (CHWs) serve between several hundred to a few thousand individuals and their catchment is generally no bigger than 10 km^2^. Due to geographic barriers in particular, systemic lack of access to health facilities for large portions of the population has resulted in community health becoming a central pillar of national health strategies globally ^6^. The lack of long-term, continuously updated surveillance datasets at the community level impedes our ability to monitor changes in disease burdens over time, locally target or evaluate the impact of community-health interventions, create outbreak detection and forecasting systems at these levels, and generally incorporate health data into decision-making processes. Given the increasing role of community programs in providing primary health care and supporting disease control efforts, the lack of routine surveillance data at this level must be remedied.

There are several barriers to the creation of a routine surveillance system at the community level. First, CHWs often only diagnose and treat common illnesses for children under 5 years old ^7^, representing only a subset of the population. Second, though officially part of national health systems, community health programs are often inadequately funded, supported, and integrated ^8,9^, with negative consequences for data completeness and quality.

For example, a case study in Malawi found that over 40% of community health reports contained errors when aggregation was conducted by CHWs due to lack of training and time available for reporting ^10^. Third, the existing structure of health system reporting often means that paper reports from the community level are submitted to district officials and integrated into the electronic HMIS system with significant delays, which limits their use for disease surveillance. An alternative is the use of health facility data disaggregated at the community level, which is becoming increasingly available with the development of new technologies such as eHealth systems. However, even when data remain disaggregated, there are issues of completeness and geographic bias due to heterogeneous access to care ^11–13^. These problems are exacerbated at fine spatial scales. For example, communities in rural areas with low access to care may be missed by routine health facility systems ^14^, significantly under-estimating disease burdens in these already vulnerable communities. Given the current lack of high-quality data at the community level, methods are needed to account for biases in these data while retaining their spatial disaggregation.

At the scale of the government health district and higher, several methods have been developed to address these issues, particularly under-ascertainment of cases (Table 1). However, none of these adjustment methods result in estimates of disease incidence that are available at the spatial scale of individual communities or at a temporal frequency that allows for rapid response. Existing methods are limited primarily by the frequency and spatial resolution of external data sources, such as large-scale surveys of disease prevalence or health-seeking behaviors. For example, information on healthcare utilization rates, such as that collected via Demographic and Health Surveys, is often collected nationally at the level of the district or region, and is inappropriate for use within smaller administrative zones. Prevalence surveys offer only a snapshot of disease burden in time, and their inferences, while available at finer spatial scales, often only apply to annual estimates. In addition, both forms of survey data are resource-intensive and are rarely available at spatial or temporal scales relevant to community health programs ^15^.

**Table 1.** Comparing the ZERO-G method to available methods for adjusting passive surveillance data.

|  | <u>Input Data</u> |  |  | <u>Output Estimates</u> |  | <u>Advantages</u> | <u>Disadvantages</u> |
| --- | --- | --- | --- | --- | --- | --- | --- |
|  | **Data Source** | **Frequency** | **Spatial Scale** | **Temporal Resolution** | **Spatial Resolution** |  |  |
| **Standard Indirect Estimators (e.g. WHO Malaria Report)** | Passive surveillance data for focal disease | Annual | Subnational (Regional) | Annual | Regional | - Straightforward adjustment method<br>- Directly accounts for health-seeking behaviors | - Only available at regional or national scales<br>- Requires adequate coverage of DHS surveys<br>- Limited to annual estimates<br>- Not appropriate for rare diseases |
|  | Survey data of health-seeking behavior (e.g. DHS) | Multi-annual | Subnational (Regional) |  |  |  |  |
| **Ecological Downscaling (e.g. Weiss et al. 2019)** | Prevalence survey | Once or Multi-Annual | Subnational (Point data) | Annual | 5x5 km | - Avoids bias in passive surveillance data | - Requires environmental and socio-economic variables<br>- Requires prevalence data with adequate spatial coverage |
|  | Environmental Variables (e.g. Bioclim) | Annual to Long-term Average | 5x5 km |  |  |  |  |
|  | Socio-economic variables | Multi-annual to annual | Regional |  |  |  |  |
| **ZERO-G Estimator** | Passive surveillance data for focal disease | Monthly | Community | Monthly | Community | - Relies solely on health system data commonly available to Ministries of Health<br>- Provides continuous, real-time estimates of incidence<br>- Corrects for missing data due to data quality issues | - Requires passive surveillance data at the community level<br>- Only appropriate for diseases with regular incidence and reporting |

Here, we introduce the zero-corrected, floating catchment gravity model estimator (ZERO-G). This method accounts for under-ascertainment of cases by public health facilities, resulting in a long-term dataset of disease incidence at the scale of individual communities or villages for common diseases that are regularly reported to the health system. Compared to existing methods, the ZERO-G estimator offers several distinct advantages for use in community health surveillance programs (Table 1). Because the main input data (notification reports and all-cause consultations) are released continuously on a set frequency, ZERO-G is able to produce estimates of disease incidence that are updated in real-time and available on a time scale relevant for decision makers. Unlike existing methods, ZERO-G relies solely on data available to local stakeholders: all-cause consultation rates, the focal disease incidence, and health facility characteristics. In addition, ZERO-G explicitly accounts for extremely low ascertainment rates that result in zero cases per month, a common occurrence in rural community health catchments. Finally, it does not rely on spatial aggregation or interpolation to combine estimates of healthcare utilization rates with disease incidence data, allowing it to retain a community-level spatial resolution.

Building on work by Hyde et al. ^16^, the method first calculates a sampling intensity derived from healthcare utilization data (i.e. consultation rates) using a floating catchment area model ^17^. It then uses spatio-temporal imputation to adjust for missing cases due to low healthcare access. This zero-adjusted data and the sampling intensity estimates are finally used to create an estimate of disease incidence that is adjusted for spatio-temporal heterogeneity in access to healthcare. This target diseases for this method are common, endemic diseases that are regularly reported to health systems in areas of high healthcare access (e.g. malaria, pneumonia, diarrheal disease). ZERO-G is not appropriate for rare diseases or those where only severe cases are reported. We demonstrate the method on a simulated endemic disease and on a case-study of field-derived passive surveillance dataset of malaria in a rural health district in southeastern Madagascar. The case study is used to further validate the ZERO-G method by comparing the estimated sampling intensity and malaria incidence rates to health- care seeking behavior and malaria prevalence from a district-representative cohort.

## THE ZERO-G ESTIMATOR

Indirect estimation methods estimate the “true” rate of disease incidence or prevalence from case data with low or uneven ascertainment rates by including information on the sampling intensity (e.g. healthcare use) in each administrative region ^18^. ZERO-G specifically combines information on the number of cases recorded by the health system with information on the proportion of cases that are expected to be observed. In addition, it includes imputation methods for adjusting for extremely low ascertainment rates that result in zero cases reported. The final result is an estimation of disease incidence equal to the expected incidence if access to healthcare was identical across space and time.

The ZERO-G estimation method can be summarized in a pseudo-statistical framework consisting of three main steps (Figure 1): 1) the estimation of healthcare access via a gravity model (Eq. 7-12), 2) the adjustment of erroneous zeroes in case notifications (Eq. 4-6) and, 3) the conversion of healthcare access to sampling intensity via multi-objective optimization (Eq. 2- 3). The estimates of sampling intensity and zero-adjusted data are then used to estimate an adjusted incidence rate (N*it)* for each administrative zone *i* and time period *t*, accounting for imperfect detection due to differing healthcare access via an Inverse Binomial distribution (Eq. 1). The full ZERO-G estimator can be stratified across demographic classes (e.g. age, sex, etc.) to account for demographically-dependent health-seeking behaviors. However, we limit our notation here to one class to improve readability. Parameters and variables representing data are further described in Table 2.

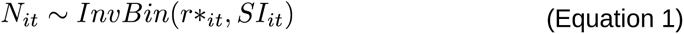

**Figure 1.**
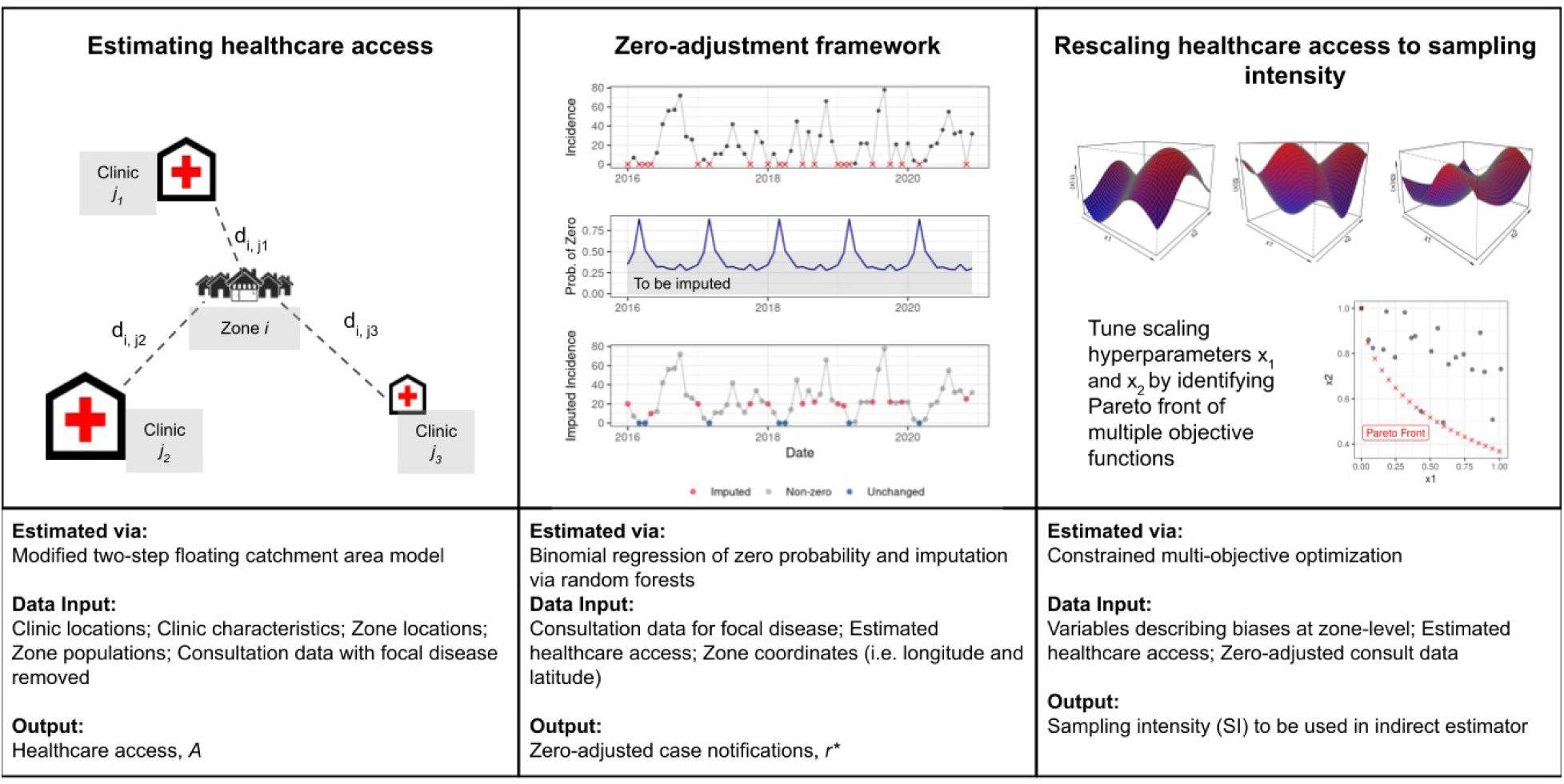
Workflow for adjusting incidence data using the floating catchment, zero- corrected (ZERO-G) estimator. Panel 1: A depiction of the gravity-model used in the floating catchment area model. A single zone *i* is represented surrounded by multiple clinics *j* with differing amount of services offered, with the distance between the zone and the clinic represented by *dij.* **Panel 2:** An example of the zero-adjustment step for one zone. Top row of Panel 2: All zeroes are identified in the dataset, represented by an X. Middle row of Panel 2: The probability of a zero is estimated via a logistic regression and those samples with a probability below 0.5 are identified. Bottom row of Panel 3: Those zeros that occur during a month with less than 0.5 probability of a zero are replaced via an imputation step. **Panel 3:** Hyperparameters are tuned via multi-objective optimization across a hyper-dimensional space, resulting in a Pareto front of non-dominated parameter values.

**Table 2.** Description of variables and parameters in ZERO-G method.

| Variable | Description | Source |
| --- | --- | --- |
| $\bar{N}_{it}$ | The ZERO-G estimated incidence after correcting for under-ascertainment | Estimated via Eq. 1 |
| $r_{it}^*$ | Zero-adjusted reported incidence | Estimated via Eq. 4 |
| $RF(X)$ | Function describing random forest algorithm used in data imputation | Estimated via Eq. 4 |
| $SI_{it}$ | The sampling intensity in zone $i$ at time $t$ | Estimated via Eq. 2 |
| $r_{it}$ | The reported incidence in zone $i$ at time $t$ | From data |
| $X_i$ | Longitude of zone $i$ | From data |
| $Y_i$ | Latitude of zone $i$ | From data |
| $m_t$ | Month of year (e.g. Jan-Dec) at time $t$ | From data |
| $\psi_{it}$ | Probability of zero reported incidence | Estimated via Eq. 5 |
| $Z_{it}$ | Binomial variable representing if there was zero reported incidence | From data |
| $\beta_{z1...zn}$ | Coefficients used to estimate probability of zero reported incidence | Estimated via Eq. 6 |
| $A_{it}$ | FCA-based healthcare access | Estimated via Eq. 7,8 |
| $h_{it}$ | Number of non-focal disease consultations | From data |
| $n_{it}$ | Population count of zone | From data |
| $g(t)$ | Function describing temporal trend in $A_{it}$. Specific function can be adjusted based on need | Estimated via. Eq. 12 |
| $S_{jt}$ | Services provided by health facility $j$ at time $t$ | Estimated via Eq. 9 |
| $C_{jt}$ | Competition at health facility $j$ at time $t$ | Estimated via. Eq. 11 |
| $v_{jst}$ | Value of health service $s$ provided at health facility $j$ at time $t$ | From data |
| $\beta_s$ | Coefficient for health service $s$ | Estimated via Eq. 9 |
| $d_{ij}$ | Distance between zone $i$ and health facility $j$. Can be calculated using Euclidean distance or based on actual routing. | From data |
| $\lambda$ | Distance decay coefficient | Estimated via Eq. 10 |
| $\beta_c$ | Scaling coefficient for competition at health facilities | Estimated via Eq. 11 |
| $x_1, x_2$ | Scaling coefficients for SI. | Estimated via Eq. 6 |

Note: Subscript $i$ refers to zone $i$ and subscript $t$ refers to time $t$ .

### Rescaling healthcare access to sampling intensity (SI) via multi-objective optimization

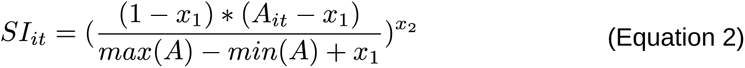

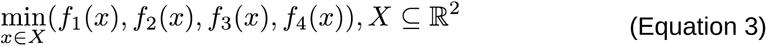

### Zero-adjustment framework

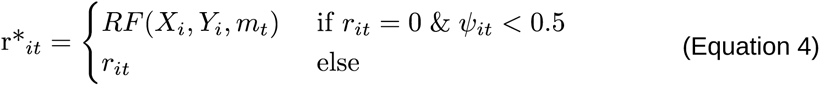

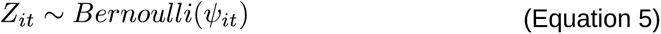

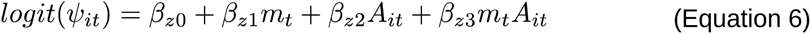

### Estimating healthcare access (A) via a gravity model

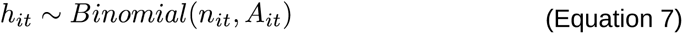

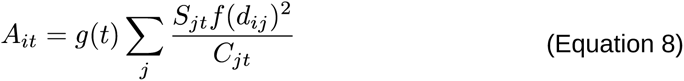

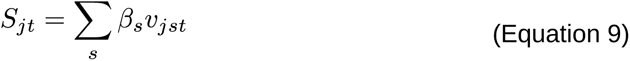

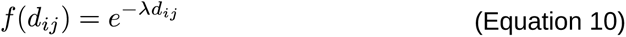

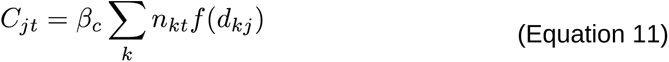

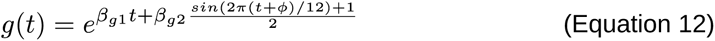

Healthcare access (A) is estimated from monthly healthcare utilization rates (i.e. consultation rates with the focal disease removed, *h_it_*). The relationship between the monthly number of consultations and the estimated healthcare access is defined via a temporally-explicit floating catchment area (FCA) model of healthcare access (Eq. 7-12) ^19^. Based on a gravity model, FCA models consider both the quantity and spatial accessibility of services at a health center for a given population by weighting the distance to care by the availability of services provided at each health center. The effect of distance on healthcare access is described by a function *f*(*d_i,j_*) that assumes exponential distance-decay, with the specific shape of the decay defined by λ (Eq. 10). Healthcare access in each community is then modeled via an “attractive force” to each health center and total access to care is the sum of these forces for a given zone (Eq. 8). Specifically, we use the modified two-step floating catchment area formulation of this metric, which allows for sub-optimal allocation of health resources via the inclusion of distance-weighted competition for each healthcare clinic’s resources ^20^. We also include a term to account for temporal trends in access due to seasonal and linear trends (Eq. 12), following Garchitorena et al. 2021^11^.

Erroneous zeroes to be adjusted are a function of both the seasonality of the disease and the estimated healthcare access of each zone. They are identified by fitting a logistic regression to the binomial variable of whether zone *i* reported zero incidence at time *t (*Eq. 4-5), resulting in estimates of the probability of a zero (ψ*_it_*). The logistic regression’s explanatory variables include the month of the year of time *t*, estimated healthcare access for zone *i* at time *t*, and the interaction between the two (Eq. 6). This logistic regression is fit to the reported case data to estimate ψ*_it_*. If zero cases are reported in a month for a zone and the probability of reporting zero cases is less than 0.5, this zero is assumed to be due to reporting error (not seasonality or low access) and is defined as erroneous. Erroneous zeros are replaced via a spatio-temporal imputation process that incorporates seasonal and spatial patterns in incidence.

Imputation is performed via 100 boosted regression tree models that estimate monthly incidence as a function of each zone’s longitude, latitude, and specific month of the zero-incidence occurrence, using the median of 100 imputations as the final imputed value (Eq. 4). Imputation is performed via the micemd package v 1.9.0 in R ^22^.

The sampling intensity (SI) is calculated from healthcare access via a constrained multi-objective optimization routine that minimizes four objective functions (Eq. 3). The objective functions correspond to: 1) the Spearman correlation coefficient between a zone’s distance to a PHC and its average annual incidence rate (geographic bias), 2) the ratio of incidence rates in zones with reimbursement policies to those without (financial bias), 3) the number of zones with annual incidence rates over 1000 cases per 1000 population (over-correction bias), and 4) the covariance of all three biases, to reduce over-correcting one value at the expense of the others. This creates a Pareto front of non-dominated values across the four objectives. From this subset, a constraint is used to limit the over-estimation of cases by constraining the results to parameters that result in monthly incidence values where the 99% percentile falls below a threshold equal to 1.5 times the original maximum monthly incidence value. The optimization routine is solved using the NSGA-II genetic algorithm via the mco package in R ^21^. The optimal values of x_1_ and x_2_ are then used to rescale Ait between x_1_ and 1 to calculate SI_it_ (Eq. 2).

## CASE STUDY: MALARIA INCIDENCE IN IFANADIANA, MADAGASCAR

We applied the ZERO-G estimator to malaria incidence in Ifanadiana District, Madagascar to demonstrate its utility in regions with highly heterogeneous rates of under-ascertainment.

Ifanadiana is a district in the Vatovavy region of southeastern Madagascar. It has an estimated population of 183,000 people spread across 195 fokontany (smallest administrative unit comprising about 1000 people) within 15 communes. Each commune contains one primary health center level 2 (PHC2), and six of the larger communes also contain a primary health center level 1 (PHC1), which provides more basic care, for a total of 21 PHCs within the district. Beginning in 2014, the Madagascar Ministry of Public Health (MMoPH) and the non-governmental organization Pivot began a partnership to strengthen the health system, establishing Ifanadiana as a model health district. This intervention works across all levels of the health system, from community health at the household level to tertiary care at the regional hospital. At the level of the PHCs, in addition to the removal of user fees, the intervention includes a range of activities to increase PHC readiness (e.g. infrastructure, equipment, supplies and personnel), support clinical programs (e.g. maternal and child health, infectious diseases), and improve data systems. As of January 2023, a minimum package of support has been been provided to all 15 PHC2s of all 15 communes, and will be expanded to a complete package at all levels of PHCs by the end of 2024. Because the progress of these health system strengthening interventions in Ifanadiana and elsewhere typically differ across PHCs and time, this requires an adjustment method that considers spatio-temporal differences in healthcare policies and interventions, such as the ZERO-G estimator.

As is common in sub-Saharan Africa^24^, the primary barriers to healthcare at PHCs in Ifanadiana are geographical and financial. The majority of the district is rural and the transportation network is primarily non-motorized; over 70% of the population lives further than an hour travel time from a PHC ^25^. As such, geographical access to care at PHCs is highly unequal, and exhibits strong distance-decay from PHC locations ^11^. Regarding financial barriers, 34% of the public health expenditure in Madagascar is out-of-pocket spending ^26^, with user fees the most cited barrier to healthcare seeking across the district ^27^. Given these known barriers, we aimed to reduce the impact of geographic and financial bias in malaria incidence rates by adjusting the data using ZERO-G.

### Data Collection

Monthly consultation data were collected at each PHC for the district of Ifanadiana from January 2016 to December 2021. Photos were taken of handwritten registries at each PHC, and patients’ residences were manually geolocated to the precision of the fokontany. The number of all-cause consultations were reported by fokontany, as well as the number of malaria cases, as confirmed by rapid detection test. Because patient ages were provided in these registries, we were able to divide the number of consultations and malaria cases into three age groups for analysis: children under 5 years old, juveniles aged 5-14, and adults aged 15 and over.

Ifanadiana suffers from shortages of diagnostic materials, specifically rapid-detection tests (RDTs) ^28^, leading to unconfirmed cases of malaria. We accounted for this reduced diagnostic capacity by scaling the confirmed malaria cases by the proportion of feverish patients who were tested via an RDT at each PHC during each month (n = 536). Information on the characteristics of each clinic by month was provided by Pivot’s Monitoring and Evaluation for Research and Learning team.

Population data came from two sources. For the 80 fokontany that receive community health program support from Pivot, we used population estimates from a Pivot-led census conducted in 2021. For the remaining 115 fokontany, population estimates came from a national census conducted in 2018 by the Madagascar National Institute of Statistics. By interpolating population values between the 2018 census and the previous 1993 census, we estimated an average annual population growth rate of 2.0%. We applied this population growth rate to both datasets to obtain each fokontany’s population by year. For both datasets, we assumed 18% of the population to be under 5 years old, 28.6% of the population to be aged 5 - 14 and the remainder to be 15 years old or above, based on the average age structure of the 80 fokontany that were censused in 2021.

Distances between residential areas and PHCs were calculated on a high-resolution transport network created via crowd-sourced mapping through a collaboration with Humanitarian OpenStreetMap. Over 20,000 km of footpaths and 100,000 buildings within the district were mapped through a two-step validation process ^25^, resulting in an open-source dataset on OpenStreetMap. Using this dataset, we estimated the distance between each household and each PHC within the district, and aggregated this to the scale of the fokontany to result in an average distance to each PHC for each fokontany. Three fokontany lacked accurate routing information and so were excluded from the analysis.

We evaluated our estimates of the SI and adjusted malaria incidence rates using external data from a longitudinal cohort survey conducted in the district of Ifanadiana (IHOPE cohort). The IHOPE cohort has conducted population-representative surveys approximately every two years from 2014-2021 using a two-stage cluster sampling scheme involving 80 spatial clusters, each containing 20 households ^29^. We include data from 2016, 2018, and 2021 in this analysis. The IHOPE cohort is based on the internationally validated Demographic and Health Surveys and is implemented by the Madagascar National Institute of Statistics. See Miller et al. ^29^ for further details on participant recruitment and study design. As part of the survey questionnaire, participants were asked if they were ill in the past four weeks and, if so, if they sought care at a public PHC. This data represented self-reported health-care seeking behavior, comparable to ZERO-G estimates of sampling intensity. Malaria prevalence data was collected via rapid detection tests conducted as part of the IHOPE survey in 2021. Briefly, children under 15 years old who consented to the study were tested for active malaria infection using SD One Step Malaria HRP-II(P.f) and pLDH(Pan) Antigen Rapid Tests. Those who tested positive were provided with a standard treatment of artesunate amodiaquine and paracetamol, with duration and dosage in accordance with national guidelines. In total, this resulted in 3774 samples across 80 clusters and 109 fokontany.

### Applying the ZERO-G estimator

#### Estimating healthcare access (A)

We estimated the healthcare access for each fokontany and month combination in our dataset following the methods described above for each age class (children, juveniles, and adults) using non-malarial consultations at PHCs. We included five traits of the health center in our calculation of Sj:

1. whether the PHC fell within the initial Pivot service catchment,
2. if point-of-care user fees (consultation costs and medications) had been removed at that time,
3. the number of staff at the PHC during each month,
4. level of health clinic (PHC1 or PHC2, with PHC2 providing more services),
5. distance from the PHC to the District office, which provides supplies, medications, and supervision.

In addition, two new PHC2 were opened in the district during the study period, one in Ampasinambo in November 2016 and one in Ambiabe in April 2018, which we accounted for in our calculation of SI. Notably, ZERO-G allows for health center traits that change over time, which we used to include monthly staffing changes, the construction of new health centers, and user fee removal interventions that were implemented over the study period.

To reduce computational time, we set a maximum limit on the distance between a community and the PHC (d_ij_) at 25 km, slightly above the maximum distance of a fokontany to the nearest PHC in Ifanadiana (22.1 km). We also included an additional parameter in our estimation of *f*(*d_ij_*) to allow the shape of this relationship to differ for those fokontany within the Pivot zone of intervention and those outside of the zone of intervention, following Garchitorena et al. ^11^

We estimated the number of non-malarial consultations *h_it_* as a random variable with a binomial distribution with the probability equal to the healthcare access (*A_it_*) and size *n_it_* equal to the population size of the fokontany (Eq. 7). Some fokontany had extremely low consultation rates and reported zero consultations for over 50% of the study period. We excluded these fokontany (n = 43) from the model fitting exercise estimating the parameters for *A_it_*, but did estimate their healthcare access from the fit model. To ensure our estimate represented the global maximum likelihood estimate (MLE), and not a local maximum, we used a two-step MLE estimation process. First, we performed a grid search via a latin hypercube sample of 1,000 samples of coarse parameter space to identify the ten parameter sets with the lowest negative log-likelihood. We then performed a second MLE step using the BFGS algorithm via the optim function in the *stats* package in R ^30^, using the parameters from the ten parameter sets with the lowest negative log-likelihood from the first step as the starting parameters. We assessed each of these ten iterations for convergence and selected the parameter set with the lowest negative log-likelihood as the optimal fit. A total of 11 parameters were estimated for each age class (Table S2.1). From the optimal parameter sets, we estimated *A_it_* for each fokontany-month combination for each age-class via Eq. 8.

#### Imputing erroneous zeroes

Nearly all fokontany (n = 189) reported zero malaria cases across all ages at least once during the study period, totaling 3468 (28.4%) of fokontany-month samples. On average, fokontany reported zero malaria cases for 18.1 months out of the 66 month period, with a range of 0 - 53 months reporting zeros. The ZERO-G method imputed between 6.08 to 10.00% of fokontany- month incidence values for each age class, an average of 4.75 months per fokontany (range: 0 – 22).

#### Rescaling healthcare access to sampling intensity

We manually set the sampling intensity (SI) to 1 for those fokontany which had an average annual healthcare utilization rate over 1 consultation per capita-year, defined as “high access fokontany” (n = 19). The remaining fokontany’s healthcare access values were rescaled following Eq. 2 and 3 using multi-objective optimization to calculate their monthly SI values.

### Evaluating Adjusted Datasets

We evaluated our estimates of the SI and adjusted malaria incidence rates using external data from the IHOPE cohort. Self-reported healthcare seeking behavior was paired spatially to SI estimates by assigning a value to a fokontany if a village from the cluster was in that fokontany.Theese data were paired temporally by taking the average of the SI during the 6 month period containing the months when the IHOPE survey was conducted in each year (January through June for 2016 and 2021 and July through December for 2018), to reduce the impact of month outliers in healthcare utilization data on SI estimates. We assessed the agreement between the two datasets by calculating the correlation between estimated SI and the proportion of residents reporting illness who attended PHCs using Clifford’s modified t-test, which controls for spatial autocorrelation ^32^. We assessed the correlation separately for each year (2016, 2018, 2021), including 109 fokontany per year.

We evaluated the ability of ZERO-G adjusted incidence rates to accurately represent malaria burdens by comparing adjusted incidence rates to malaria prevalence data collected via the IHOPE cohort in 2021. The two datasets were paired spatially by assigning a value to a fokontany if a village from the cluster was in that fokontany and were paired overtime by matching the month of the IHOPE survey to the month of the incidence rates. Because the relationship between prevalence and incidence is non-linear, we transformed cluster-level prevalence rates into incidence rates following a previously published model^33^ to allow us to compare incidence rates from both datasets. However, there remain important differences between this measure of incidence and that derived from case notifications. Prevalence data may under-estimate malaria incidence as the conversion results only in symptomatic cases of malaria while case notifications may include a higher proportion of asymptomatic cases due to co-infection with a second febrile-inducing pathogen ^34^. We compared adjusted incidence rates for children under 15 years old to prevalence rates of children under 15 years old from the IHOPE cohort for all fokontany with information in both datasets (n= 109) via Clifford’s modified t-test. We also assessed the ability of the adjusted incidence data to correctly identify hot spots of malaria, defined as the quartile of fokontany with the highest prevalence values.

## APPLYING ZERO-G TO A SIMULATED DISEASE

To demonstrate its generalization, we used the ZERO-G estimator to adjust for under-ascertainment of cases of a simulated endemic, seasonal disease. We simulated a model health district containing 100 administrative zones and 8 health clinics that differed in the number of staff, whether they offered advanced services, and whether health care was subsidized. We then simulated disease dynamics for a constant background disease rate and for two additional diseases that exhibited annual seasonality for each administrative zone at a monthly frequency for five years. We modeled an individual’s probability of seeking care as a random variable with probability equal to that zone’s reporting rate, itself a function of its distance to a clinic and the services available at that clinic, plus a random error term (Eq. S2). To represent realistic issues in data quality, we also simulated months reporting zero cases as both a function of low reporting rates and low disease incidence and due to randomness. This resulted in a time series of “true” disease incidence and reported disease incidence for each zone over a five year period (Fig. S3). Further details on the creation of the simulated dataset are reported in the Supplemental Materials.

The performance of the ZERO-G method on the simulated data was evaluated by comparing the ability of the ZERO-G to reproduce the original simulated “true” data. We calculated the root mean squared error (RMSE) and correlation coefficient between the true incidence and adjusted incidence rates across patches and seasons. We compared these values to the unadjusted incidence rates to assess the improvement provided by the ZERO-G method.

## ETHICS STATEMENT

Use of aggregate monthly healthcare utilization data from PHCs in Ifanadiana District for this study was authorized by the Medical Inspector of Ifanadiana. The IHOPE longitudinal survey implemented informed consent procedures approved by the Madagascar National Ethics Committee and the Madagascar Institute of Statistics. Household-level de-identified data from the IHOPE survey were provided to the authors for the current study. We recognize that all research is conducted within the surrounding socio-political context and risks reproducing existing inequalities within the research team and across research partners. We’ve chosen to explicitly reflect on power dynamics and equitable authorship within the context of this research project in an accompanying reflexivity statement (Supplemental Materials).

## RESULTS

### Case Study: Malaria in Ifanadiana, Madagascar

We estimated the SI by fitting a floating catchment area model to healthcare utilization data from January 2016 - December 2021 and rescaling it via multi-objective optimization. The resulting model performed well at reproducing the healthcare utilization data (under-5: Spearman’s ρ = 0.619, p-value<0.001; juvenile: Spearman’s ρ = 0.608, p-value<0.001; adult: Spearman’s ρ = 0.702, p-value<0.001). When averaged over all fokontany per month, it accurately represented the temporal trends in the healthcare utilization data, although this performance was dependent on age-class (under-5: Spearman’s ρ = 0.384, p-value <0.01; juvenile: Spearman’s ρ = 0.517, p-value <0.001; adult: Spearman’s ρ = 0.578, p-value < 0.001). When averaged across time to result in one average SI per fokontany, it also was able to capture spatial and fokontany-specific differences in healthcare utilization rates (under-5: Spearman’s ρ =0.829, p-value <0.001; juvenile: Spearman’s ρ =0.806, p-value <0.001; adult: Spearman’s ρ =0.844, p-value <0.001).

The spatial patterns in the estimated SI mirrored spatial patterns in self-reported healthcare seeking behavior from the IHOPE longitudinal survey (Fig. 2). The estimated SI and self-reported healthcare seeking rates were significantly correlated across all years (Clifford’s t- test; 2016: ρ = 0.502 (p < 0.01), 2018: ρ = 0.644 (p < 0.01), 2021: ρ = 0.564 (p < 0.01), Fig S2.1). Both data sources estimate higher healthcare access at fokontany nearer the national transportation network, specifically the paved road that runs east-west through the district, and in close proximity to PHCs. In addition, the two datasets were in agreement that the majority of the district has low access to healthcare.

**Figure 2.**
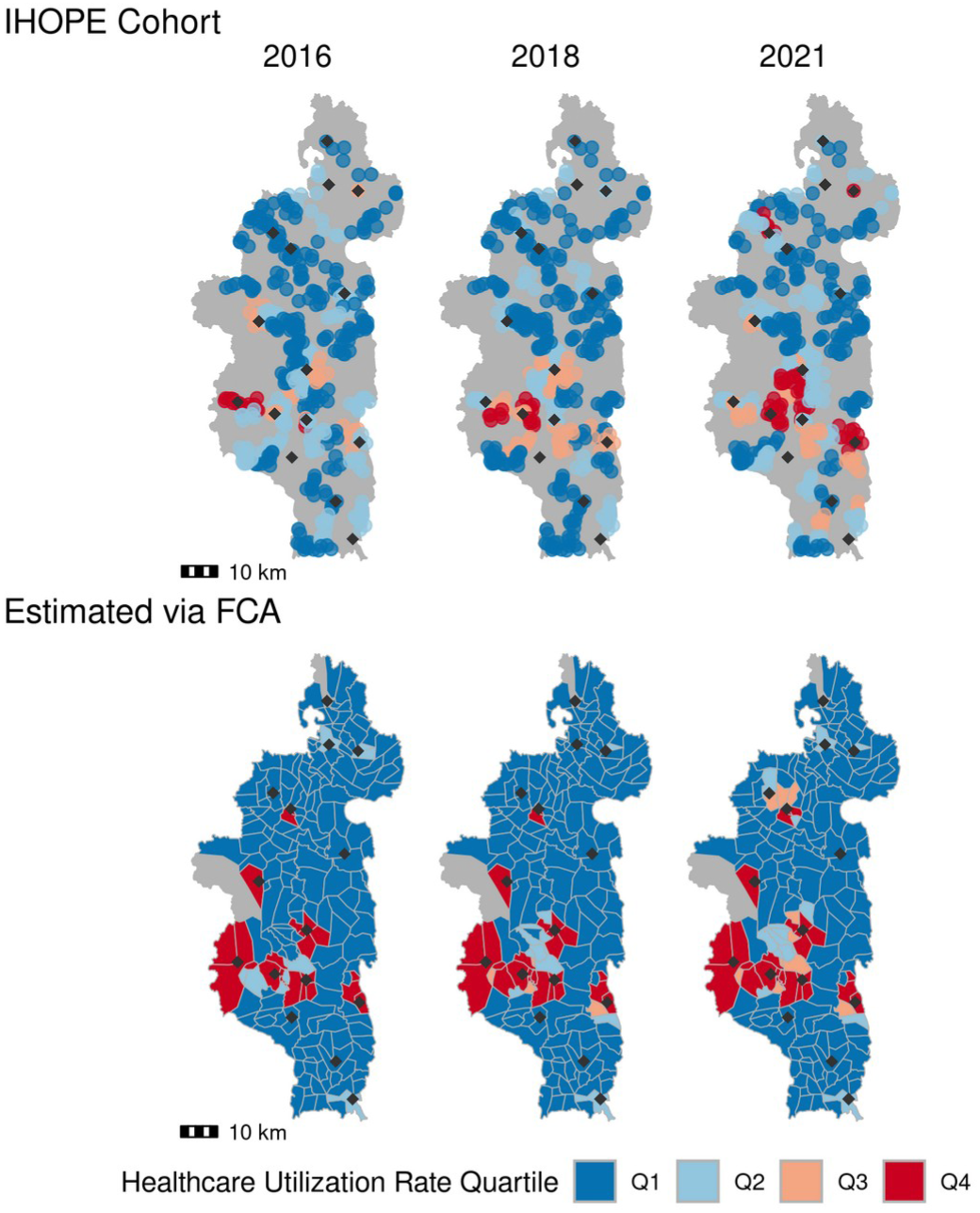
The sampling intensity estimated via the gravity model and multi-objective optimization (bottom row) closely approximates self-reported healthcare seeking rates from the IHOPE cohort (top row). Shading represents rates grouped into quartiles, with Q1 corresponding to the lowest healthcare utilization rate and Q4 to the highest. Diamond points show the location of level-2 PHCs. Top row: Cluster-level healthcare seeking rates are illustrated for each village in a cluster across the three survey years. Bottom row: The scaled sampling intensity estimated via Eq. 2-3 & 7-12. Scatter plots of this data are shown in Fig S2.1

### Reduction of Bias in Malaria Incidence due to Geographic and Financial Barriers to Care

The unadjusted dataset showed evidence of geographic bias; average annual incidence of malaria in a fokontany was negatively correlated with the distance from that fokontany to the nearest PHC (Spearman’s ρ = -0.617, p-value < 0.001, Fig. 3A), showing an exponential distance decay. The adjusted dataset, by comparison, demonstrated no relationship between average annual incidence and distance to the nearest PHC (Spearman’s ρ = -0.060, p-value = 0.409, Fig. 3A). Fokontany whose populations attended PHCs where fees were removed for the user (PHCs were reimbursed by Pivot) reported 2.48 times higher incidence than those that did not benefit from the reimbursement policy in the unadjusted dataset (Fig. 3B). Applying the ZERO-G method drastically reduced this bias; the average annual incidence in these fokontany was 0.95 times the incidence in fokontany with cost-of-care-reimbursement (Fig. 3B). However, this reduction in bias differed across years. Specifically, zones with reimbursement policies retained a much higher incidence rate in 2018. This difference was driven primarily by high monthly incidence (>500) in the unadjusted data due to a malaria outbreak in the north of the district in a commune benefiting from fee-reimbursement. Because it does not aggregate or smooth incidence data, ZERO-G retained this anomaly in incidence rates even after adjustment. This is an advantage of ZERO-G, as it allows for the identification of epidemics or unexpected trends in the data.

**Figure 3.**
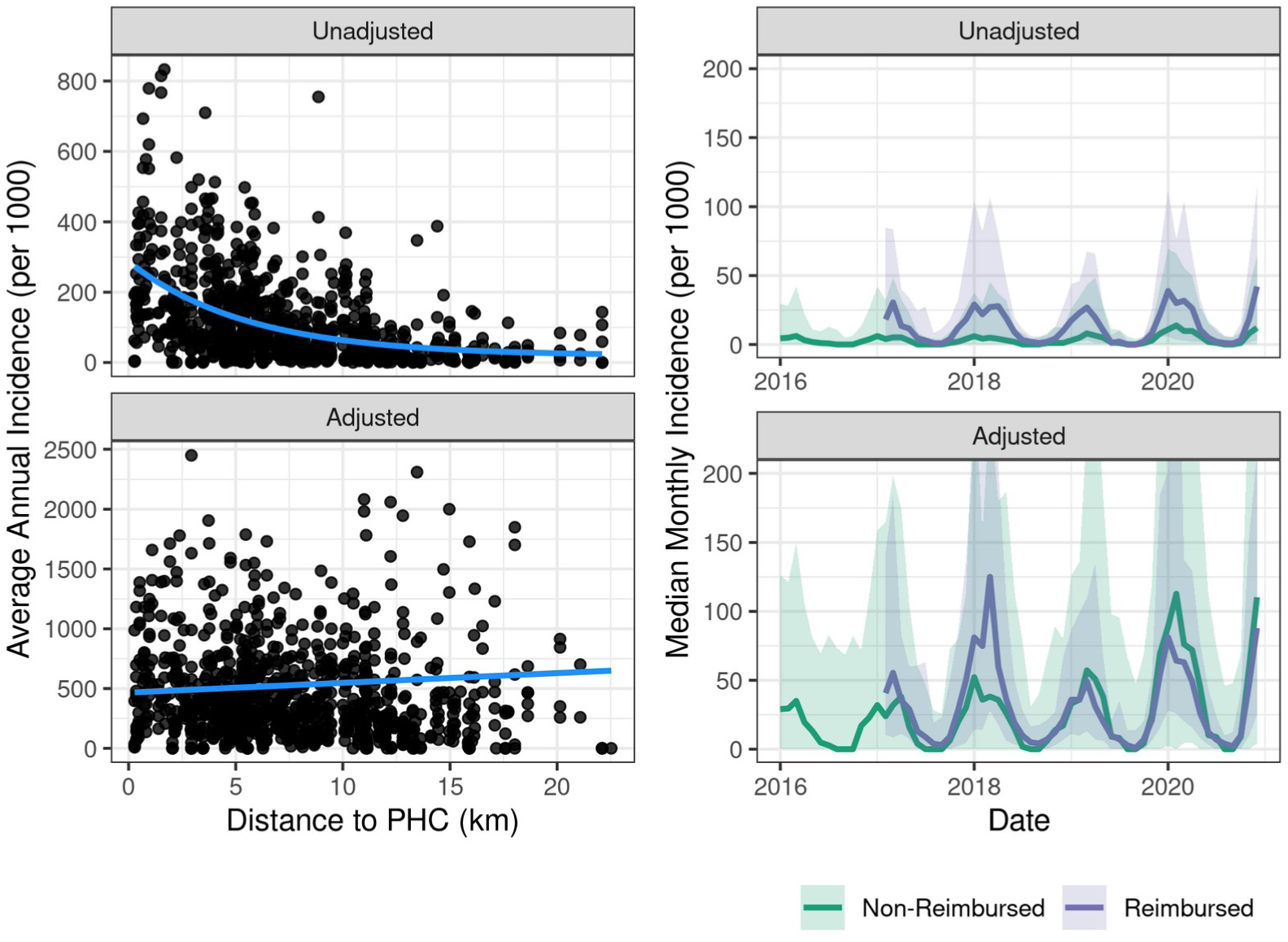
The ZERO-G adjustment method greatly reduces geographical and financial bias in malaria incidence rates. Left: Each point represents the average annual malaria incidence rates for a fokontany over the period of 2016-2020, with the x-axis showing the distance to the nearest PHC. The smoothed line is the exponential (unadjusted) or linear (adjusted) fit between average annual incidence and distance to PHC. One outlier point is removed to aid with visualization. Right: The median monthly malaria incidence rates across fokontany whose closest PHC does or does not offer fee reimbursement. Fee reimbursement began in January 2017. The error ribbon represents a 90% CI. The y-axis is limited between values of 0-200 to aid with visualization.

### Comparing Unadjusted and Adjusted Datasets

Comparing the unadjusted and adjusted datasets, we estimated that unadjusted case notifications are capturing on average 26.5% of symptomatic malaria cases in the district. This differed by year, with the lowest percentage of 24.2% in 2016 and the highest of 31.6% in 2017. The level of under-ascertainment also varied across fokontany. On average, the adjusted annual incidence in a fokontany was 9.15 (range: 1- 451) times the unadjusted annual incidence rate. However, when this was calculated omitting fokontany and year combinations that reported zero malaria cases in a year (26 out of 944), this ratio was reduced to 8.42 (range: 1 – 76.5).

### Validation with Prevalence Data

We validated ZERO-G by comparing ZERO-G estimated incidence rates with incidence rates derived from the IHOPE prevalence survey in children under 15 years old (Fig. 4). Unadjusted incidence rates were negatively correlated with IHOPE incidence rates based on prevalence, but this correlation was not significant (Spearman’s ρ = -0.141, p-value = 0.2). The unadjusted incidence rates had no correlation with the calculated incidence of symptomatic individuals in the IHOPE survey (Spearman’s ρ = -0.050, p-value = 0.6). After adjusting the data, we found a positive correlation between ZERO-G and IHOPE incidence rates (Spearman’s ρ = 0.316, p-value = 0.001). While the estimated correlation coefficient between incidence rates and the proportion of symptomatic and RDT positive children was positive in the adjusted dataset, it remained insignificant (Spearman’s ρ = 0.188, p-value = 0.06). The adjusted dataset also doubled the number of correctly-ranked fokontany into quantiles that matched those from the prevalence data (Fig. 4). The adjusted dataset correctly ranked 43 of 104 fokontany, compared to 18 in the unadjusted dataset.

**Figure 4.**
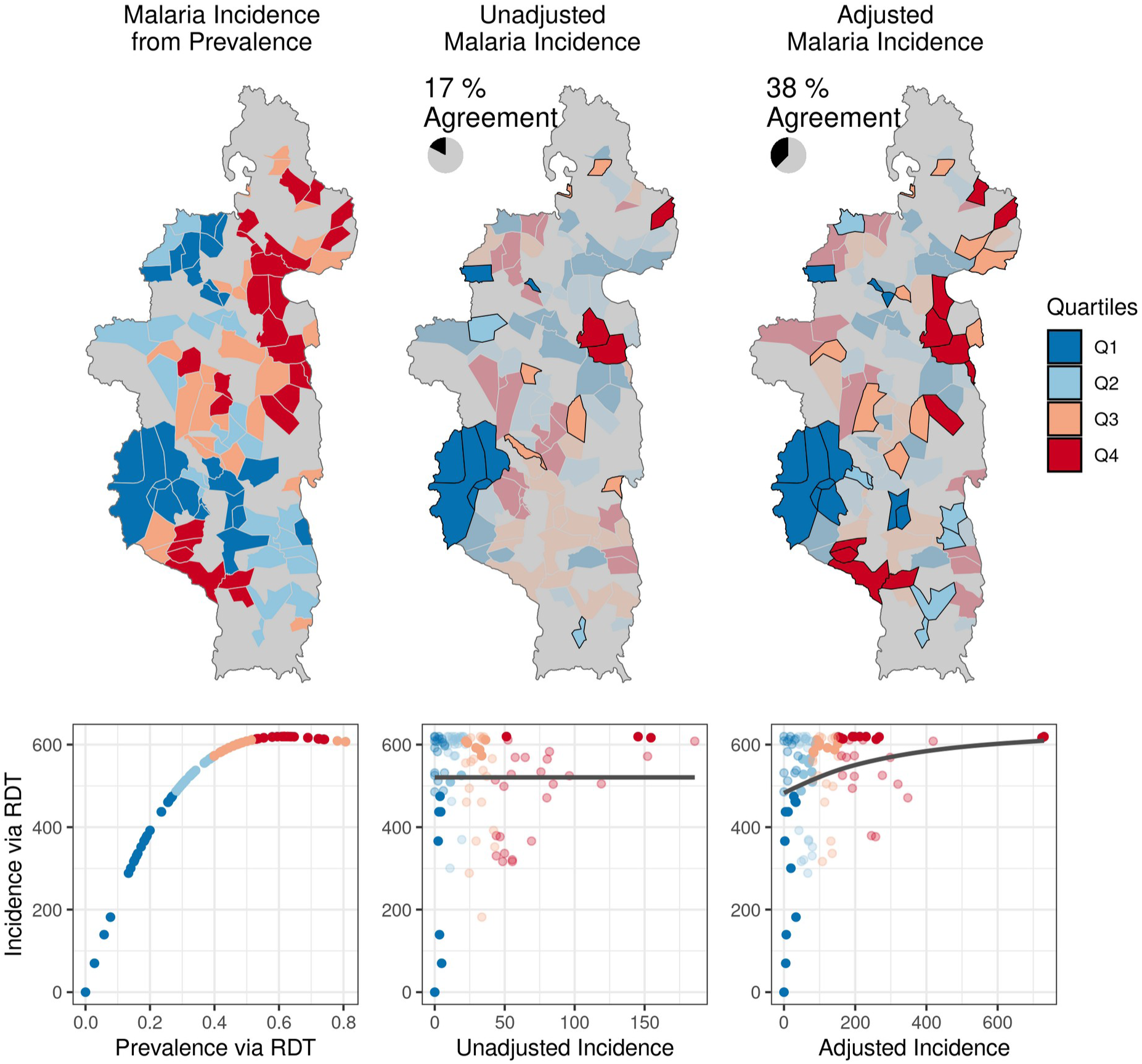
The adjustment method results in monthly malaria incidence rates in 2021 that more closely correspond to measures of malaria prevalence in children under 15 years old. Left: Malaria incidence derived from prevalence as measured by rapid-detection tests (RDT) in children under 15 years old from the IHOPE cohort survey. Colors represent quartiles from Q1 (lowest incidence) to Q4 (highest incidence). The scatter plot illustrates the non-linear relationship between prevalence and incidence. Middle: Monthly malaria incidence in the unadjusted dataset. Quartiles that match those in the prevalence data are highlighted in black. The scatter plot illustrates the relationship between unadjusted incidence and IHOPE incidence. Right: Monthly malaria incidence in the ZERO-G adjusted dataset. Quartiles that match those in the prevalence data are highlighted in black. The scatter plot illustrates the relationship between ZERO-G incidence and IHOPE incidence. Monthly incidence has been chosen to correspond to the month in which the IHOPE survey was conducted for that fokontany.

### Simulated Endemic Disease Data

The ZERO-G estimator reproduced simulated true incidence data when applied to simulated reported incidence data that contained reporting biases due to healthcare access. The ZERO-G adjusted dataset predicted the true monthly incidence rates with a RMSE of 54.5, compared to a RMSE of 94.67 when using the unadjusted dataset (Fig. S1.4). It was also more strongly correlated with the true incidence rates (Spearman’s *ρ* =0.82, p-value<0.001), compared to the unadjusted dataset (Spearman’s *ρ* =0.43, p-value<0.001) (Fig. S1.5). In addition, the ZERO-G adjusted dataset reduced biases due to geographic distance and fee reimbursement policies seen in the unadjusted dataset (Fig S1.6, Fig S1.7). The unadjusted incidence dataset exhibited a strongly negative correlation with increasing distance to the nearest health clinic (Spearman’s *ρ* = -0.666, p-value <0.001), which was reduced by over 30% in the ZERO-G adjusted dataset (Spearman’s *ρ* = -0.465, p-value<0.001). The ratio of incidence in zones served by health clinics offering fee reimbursement to incidence in zones without this policy was only 1.11 in the ZERO-G adjusted dataset, compared to 1.93 in the reported dataset and 1.00 in the true dataset.

## DISCUSSION

There is a critical need for routine surveillance systems to produce estimates at the spatial scale of individual communities so that control interventions can be targeted in collaboration with community health programs. However, HMIS data are rarely kept disaggregated at this scale and, when they are, they suffer from under-estimation of incidence that varies across space and time, preventing their usefulness for decision making . We developed an adjustment method that combines a gravity-model of healthcare access with an indirect estimator to create long-term routine surveillance data at the community-scale, adjusted for under-ascertainment due to uneven health care access. We demonstrated this method by applying it to field-collected malaria case notification data from 192 communities over 5 years of surveillance in a rural District of Madagascar. This method reduced geographical and financial bias in field-collected malaria incidence rates by 91% and 96%, respectively. In addition, we validated this method with two external, population-representative datasets and found strong agreement with self-reported healthcare access and malaria prevalence rates. We further assessed the generalizability of the ZERO-G estimator on a simulated dataset and found it nearly doubled the ability to reproduce true incidence rates. The ZERO-G estimator can obtain estimates that approximate long-term active surveillance data of common, endemic diseases at fine-spatial scales using only passive surveillance data.

ZERO-G greatly reduced bias in malaria incidence rates from a passive surveillance dataset in our case study. In Ifanadiana district, per capita health system utilization rates are twice as high for fokontany within 5km of a health center than those further away ^11^, and we found similar trends in the unadjusted malaria data (Fig. 3). Geographic bias in the malaria data was therefore primarily reduced by accounting for low sampling intensity at those fokontany further than 5 km from a PHC (Fig. 2). Financial costs represent a significant barrier to healthcare seeking, particularly for low-income communities, and differential user fee policies over time (e.g. implementation of universal health coverage) can result in healthcare access patterns changing as a function of this ^35,36^. In Ifanadiana, the removal of user fees to patients (via reimbursement policies to PHCs) in part of the district led to a sudden and sustained 65% increase in utilization rates ^27^. ZERO-G removed this bias, resulting in similar incidence rates regardless of when and where reimbursement policies were in place. ZERO-G also resulted in data that more accurately identified malaria prevalence hotspots and coldspots than the unadjusted data, performing twice as well. However, the adjusted dataset only correctly categorized 38% of fokontany into ranked quantiles, illustrating the difficulty in matching incidence data to prevalence data. While we accounted for the non-linear relationship between malaria incidence and prevalence in our evaluation of ZERO-G, we did not account for age-specific differences in symptomatic rates between children and juveniles ^37^, which may have further skewed this comparison. Further, we only had access to one study of malaria prevalence at a spatial-scale finer than 5 x 5 km. Therefore, we were only able to assess our method’s ability to reproduce spatial patterns in malaria burden, and not temporal patterns. However, our model results agree with national-level trends in malaria, which witnessed over a 40% increase in confirmed malaria cases in 2020 ^38^, suggesting we are capturing temporal trends as well.

Unlike other methods, which rely on external datasets describing sampling intensity that are collected at coarse spatial resolutions and infrequently (e.g. DHS, MICS, or other survey data), ZERO-G uses data that match the spatial and temporal resolution of the case notification data. This allows it to retain the original spatial and temporal scales at which the data was collected while relying solely on public health and demographic data that is easily accessible to public health actors. Population data can be sourced at fine-scale administrative levels via national census data or via open-source datasets such as PopGrid ^39^. As with all estimates of population-level indicators, the lack of high-quality population estimates (the “denominator problem”^40^) is an obstacle to estimating incidence rates and may lead to biased estimates.

Information on PHC locations and services are collected by Ministries of Health or available via regional, open-source datasets (e.g. ^41^). These data may not always be available on a monthly basis, particularly staffing data. In these cases, annual or static data may be substituted for monthly data, as demonstrated in the Madagascar case study. In the context of health interventions, however, the ability to track monthly changes to policies or health infrastructure due to an external intervention is a benefit of the ZERO-G estimator over existing methods. We used a field-verified transport network created via OpenStreetMap to estimate the distance between a population and a PHC, which accurately represents patients’ distance to PHCs ^25^; however, these transportation networks are not globally available. When transportation networks are not available, open-source databases of populations’ distances to PHCs and other services could serve as suitable substitutes (e.g. ^42,43^). Finally, consultation rates are commonly tracked by health systems and are increasingly recorded via electronic health management information systems ^44,45^, facilitating their use in these estimates.

ZERO-G differs from existing adjustment methods in several ways. First, it uses monthly estimates of sampling intensity in the indirect estimate step rather than data from annual or inter-annual population surveys. Most adjustment methods do not account for changes in healthcare seeking behavior due to seasonality or temporal shifts to the health system (e.g. climate-driven changes in access, changes in PHC staffing rates, clinic-level interventions), and are therefore limited to inference at an annual frequency ^46^. This functionality of the ZERO-G method is particularly beneficial in the context of partial health system interventions, such as the adoption of new policies or technologies. Second, the resulting dataset is available at the same spatial scale at which it is collected, rather than spatially interpolated between points or aggregated to coarser resolutions. We build on work by Hyde et al. ^16^, which proposed a similar indirect estimation adjustment method for malaria data that featured a monthly frequency at the scale of the community, but dealt with extreme low incidence values by spatially smoothing estimates between neighboring communities, introducing spatial structure into the adjusted dataset and removing existing natural variation. Because ZERO-G estimates are available at the community level at a monthly frequency, they can be used to inform community health programs and spatially targeted interventions at the village level in real-time, capabilities that are lacking in other adjustment methods. In addition, ZERO-G explicitly models the sampling intensity as a function of geographic and health-system characteristics in all the facilities surrounding a community via a gravity model instead of using information from the closest facility in a linear model, as in Hyde et al. ^16^. Because of this, changes in the health system, such as the closing of a facility due to a natural disaster or a policy change, can be directly incorporated into calculations of sampling intensity in near real-time. It also allows for estimation of sampling intensity in unsampled communities or months through these modeled processes, rather than relying on interpolation.

There are several limitations that should be taken into consideration when implementing ZERO-G. First, the adjustment of zero-incidence samples due to extremely low ascertainment introduces a further source of uncertainty. However, the identification of which samples to impute is data-driven, and, as demonstrated when applied to both the simulated and field-derived datasets, replaces only a small fraction of the overall data. Secondly, the ZERO-G estimator does not include a step to disaggregate consultation rates to a finer spatial scale than that reported by the PHC, often a major limiting step to accessing disease incidence data at a fine spatial scale. In Ifanadiana, the standard reporting system aggregates consultations at the level of the health facility catchment. We manually digitized health registers to obtain community-level data, a time- and resource-intensive process. However, the increased availability of electronic systems at the level of primary and community health care represents an opportunity to apply this method directly and in real time to data at fine spatial scales. Finally, the ZERO-G method is not appropriate for all passive case notification datasets. It is best suited for routine passive surveillance of common, endemic diseases, which possess the historical datasets needed to impute low-incidence values. The ZERO-G method is also inappropriate for adjusting case notifications of novel diseases because behavioral and health-system responses to a rapidly-evolving epidemic will violate the assumption that the relationship between healthcare access and sampling intensity of the disease is constant.

In conclusion, ZERO-G represents a promising new method for adjusting passive surveillance data of endemic diseases for under-ascertainment bias in regions with low and heterogeneous healthcare seeking rates, developed specifically for use at the community level. Unlike other methods, it is applicable in regions with ongoing heterogeneous public health interventions, allowing it to be used to adjust case notifications used in monitoring and evaluation efforts in addition to routine monitoring of diseases. This method can serve as part of a wider toolkit of statistical techniques used to improve targeted health system responses. In a case study in a rural health district in Madagascar, it was able to reduce geographic and financial bias in malaria incidence and the resulting dataset more closely approximated spatial trends in malaria prevalence. It is particularly suited to rural areas, where geographic isolation strongly influences healthcare access ^42^. As spatially-explicit health metrics become an increasingly important tool for precision public health interventions, there is an urgent need to obtain and use quality data sources at the community scale. Statistical methods such as ZERO-G can be an important tool to support the role of community health programs in the local targeting of interventions for disease control.

## Data Availability

All code and data needed to reproduce this study are available in a figshare repository (doi: 10.6084/m9.figshare.22154492).

https://doi.org/10.6084/m9.figshare.22154492.v1

## ACKNOWLEDGMENTS

We would like to thank the health professionals who collect passive surveillance data in addition to serving their patients and the Pivot data collection team for their work collecting and digitizing this data. We would also like to thank Ann Miller and Marius Randriamanambintsoa for their support of the IHOPE longitudinal survey.

