## Supplemental Materials for "The Zero-Corrected, Gravity-Model Estimator (ZERO-G): A novel method to create high-quality, continuous incidence estimates at the community-scale from passive surveillance data"

### Table of Contents

1. Creating Simulated Incidence Data
2. Case Study: Malaria Incidence in Ifanadiana, Madagascar
3. Reflexivity Statement

### 1. Creating Simulated Incidence Data

*Spatial distribution of health care infrastructure*

Monthly disease incidence was simulated for 100 administrative zones (patches, *p*) over 5 years. The patches were distributed in a 10 x 10 square matrix representing a health district (Figure S1.1). Each patch's population was drawn from a uniform distribution between 800 and 1200 and age-stratification was not considered. The population remained constant over the simulated time period.

Eight primary health clinics (*j*) were randomly distributed across the 10 x 10 matrix (Figure S1). Clinics differed in the number of staff (random uniform from 5 to 15), whether they offered advanced services (randomly distributed so that 50% of clinics offered advanced services), and whether health care was provided free-of-charge at the clinic (randomly distributed so that 50% of both advanced and basic clinics offered this service).

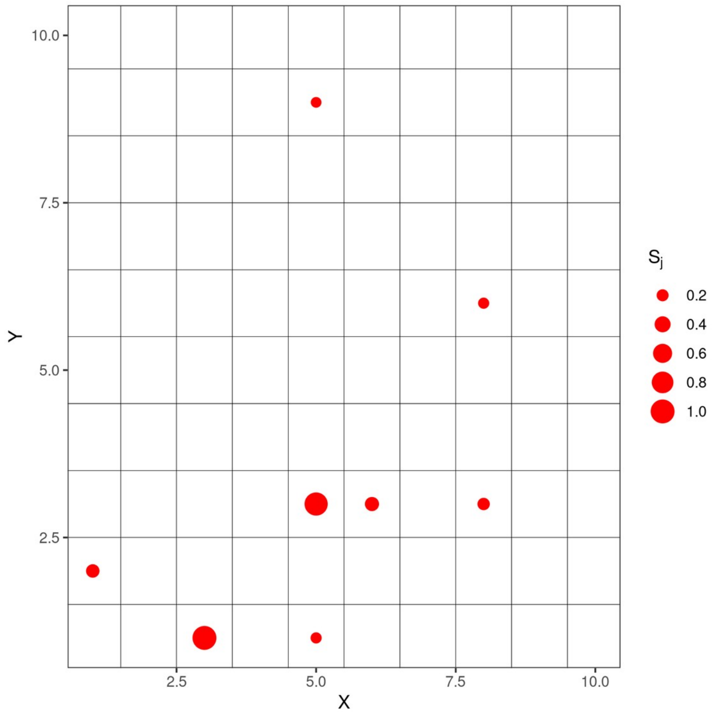


**Figure S1.1. Distribution of primary health clinics (red points) distributed among a matrix of 10 x 10 administrative zones (squares).** The size of the point for each clinic corresponds to the level of services it provides ($S_j$).

*Disease Dynamics*

We simulated consultation rates for constant background disease rates and for two diseases that exhibited annual seasonality in their burdens. We assumed the background disease rate was one infection per person per year. We set the annual incidence of each seasonal disease to one infection per person per year, but varied the seasonality of each disease separately, resulting in a monthly risk of infection for each disease *g* ($\phi_g$) (Figure S1.2). Each individual's probability of infection for each disease during each month was defined as the inverse logit of the logit-transformed monthly risk of infection plus a normally distributed random error with a mean of 0 and standard deviation of 0.5, resulting in a probability ranging from 0 - 1. This extra error was added so that the simulated data approximated the noisiness seen in field-derived disease notification data. This resulted in a number of cases for each disease *g* in patch *p* during month *t* drawn from a binomial distribution of size equal to the patch's population and probability $\phi_g$ (Eq. S1).

$$C_{g,p,t} = Bin(population_p, \phi_g) \qquad \text{(Equation S1)}$$

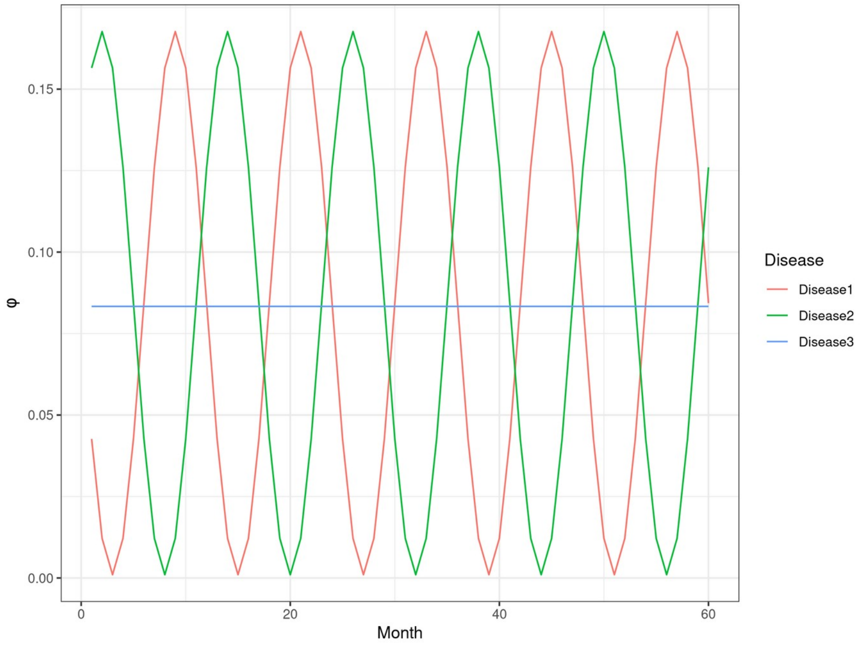


**Figure S1.2. The monthly risk of infection (phi) for each of three diseases across the simulated time period.**

*Reporting Rate*

We modeled an individual's probability of seeking health care at the patch level ($PC_p$) as a function of the distance to health clinics and the characteristics of those clinics (Eq. S2).

$$PC_p = \sum_j S_j(e^{-0.3d_{pj}^2}) \qquad \text{(Equation S2)}$$

Where $\mathrm{S}_j$ is the services provided by each clinic j and $d_{pj}$ is the distance between patch *p* and clinic *j*. The services provided by each clinic *j* were a function of the number of staff of that clinic $(\mathrm{x1})$, whether it offered advanced services $(\mathrm{x2})$, and whether healthcare was provided free of charge $(\mathrm{x3})$, scaled to range from 0 – 1 (Eq. S3).

$$Sj = \frac{x1_j * (x2+1) * (x3+1)}{\max_{k\in[j]} S_j} \qquad \text{(Equation S3)}$$

In addition, we simulated instances of zero reported infections per patch for each disease due to 1) a combination of low reporting rates and low disease risk and 2) randomness. These two causes of zero reported infections were simulated independently from each other by a random binomial event given a corresponding probability of a zero. The probability of a zero due to low reporting rates ($PC_p$) and low disease risk ($\phi_g$) for each disease *g* in patch *p* at month *t* was calculated following Equation S4:

$$Pz_{g,p,t} = 1 - PC_p^{1-\phi_g^{0.1}} \qquad \text{(Equation S4)}$$

The probability of a zero due to randomness (Pzr) was set at a constant value of 0.1.

The number of reported monthly cases for each disease per patch was therefore defined as:

$$R_{g,p,t} = Bin(C_{g,p,t}, PC_p) * Bin(1,(1 - Pz_{g,p,t}) * Bin(1, 1 - Pzr) \qquad \text{(Equation S5)}$$

An example of comparing the true vs. reported cases for all three diseases for two patches of differing health care access is shown in Figure S1.3. Notably, this dataset bears characteristics that resemble realistic passive notification datasets, including a high variance around the mean and unexplained missing data reported as zeros.

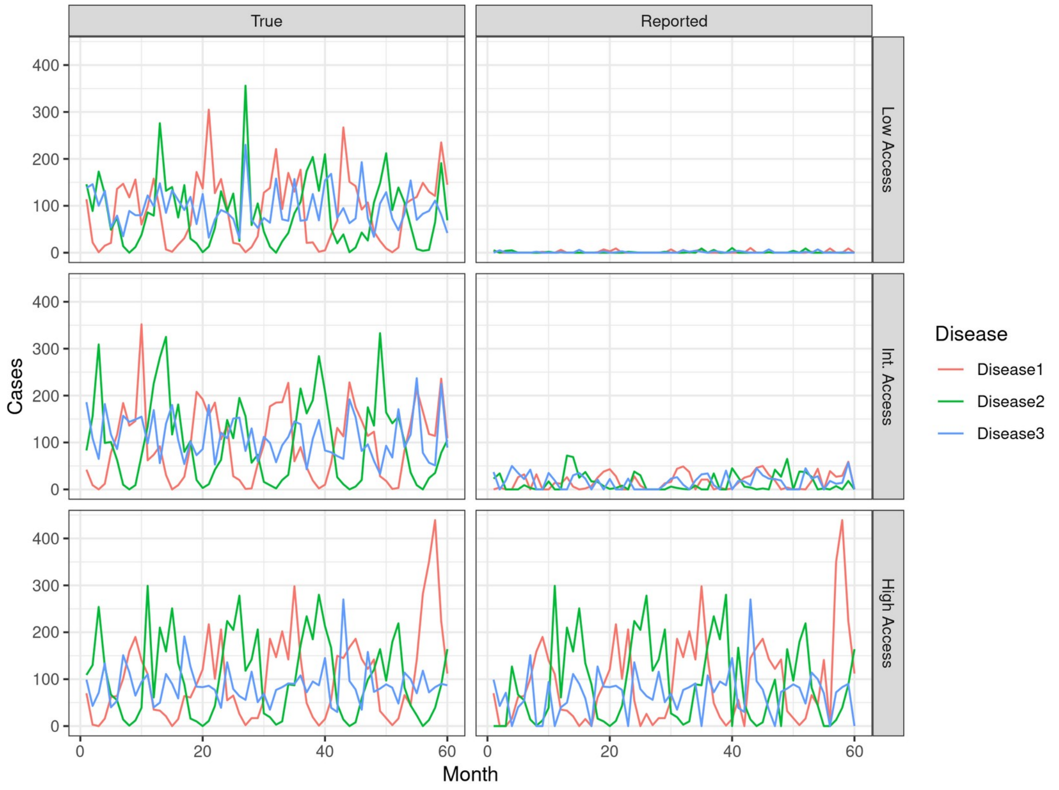


**Figure S1.3.** The true number of cases and reported number of cases for all three diseases in three example patches with low, intermediate, and high probability of seeking healthcare.

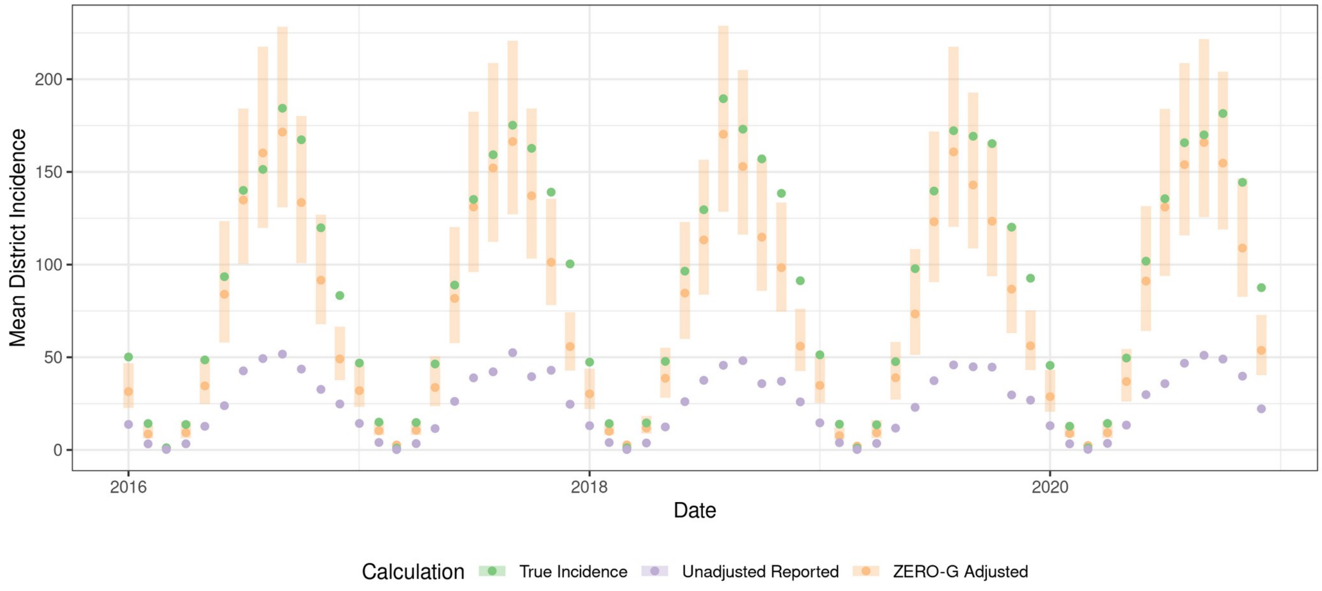


**Figure S1.4.** Time series of district-level incidence rates in the simulated dataset, for the true dataset, reported dataset, and ZERO-G adjusted dataset. The bars represent the 95% confidence intervals of the ZERO-G adjustment.

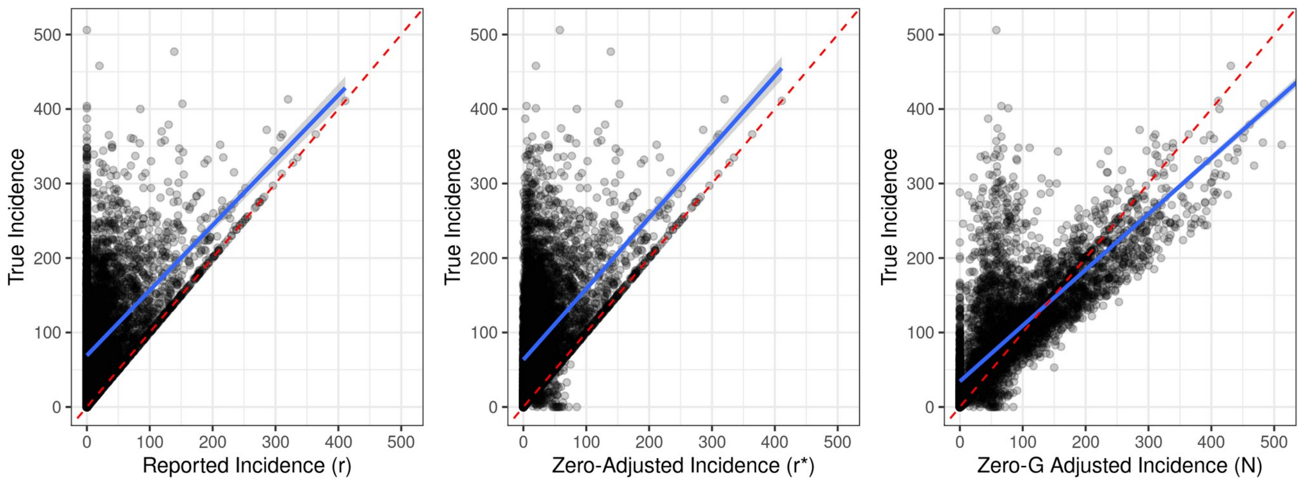


**Figure S1.5** Scatter plots comparing simulated true incidence data with unadjusted incidence rates, incidence rates adjusted for erroneous zeros, and ZERO-G adjusted incidence estimates.

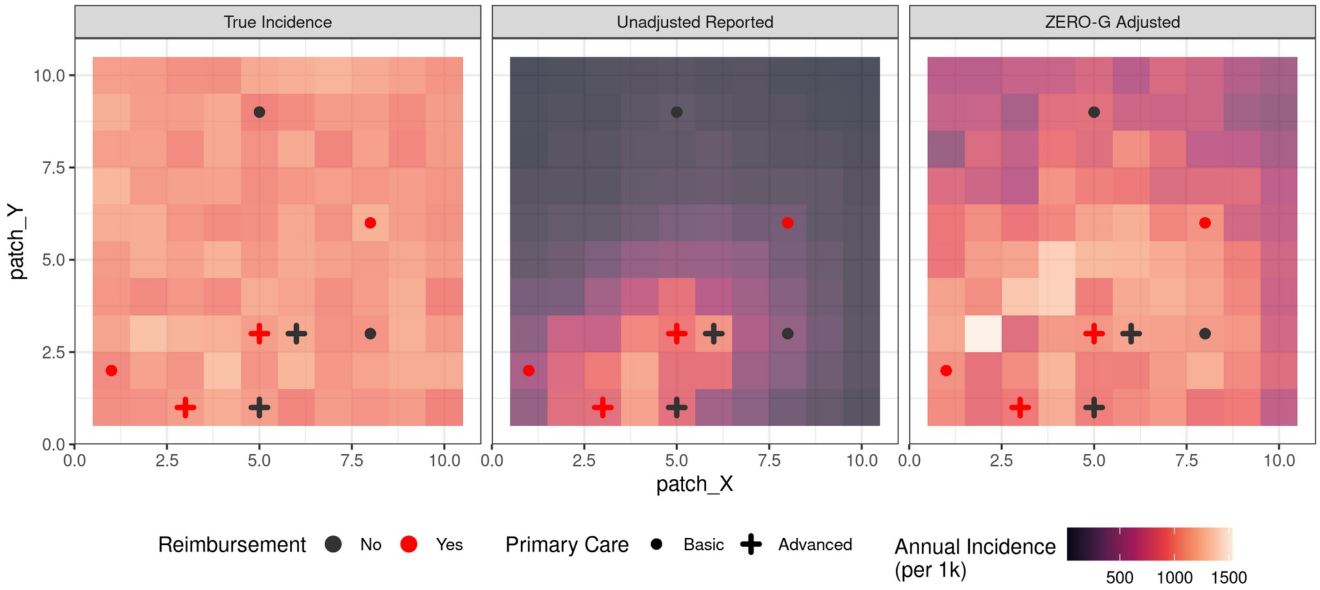


**Figure S1.6.** Spatial pattern of mean annual incidence per administrative zone in the true incidence dataset, reported dataset, and the ZERO-G adjusted dataset. The annual incidence is represented by the shaded color of each zone and the location of health clinics are represented by the points. Health clinics offering advanced primary care are represented by a cross and those offering basic care are represented by a circle, with the color of the point corresponding to whether fees are reimbursed at that clinic.

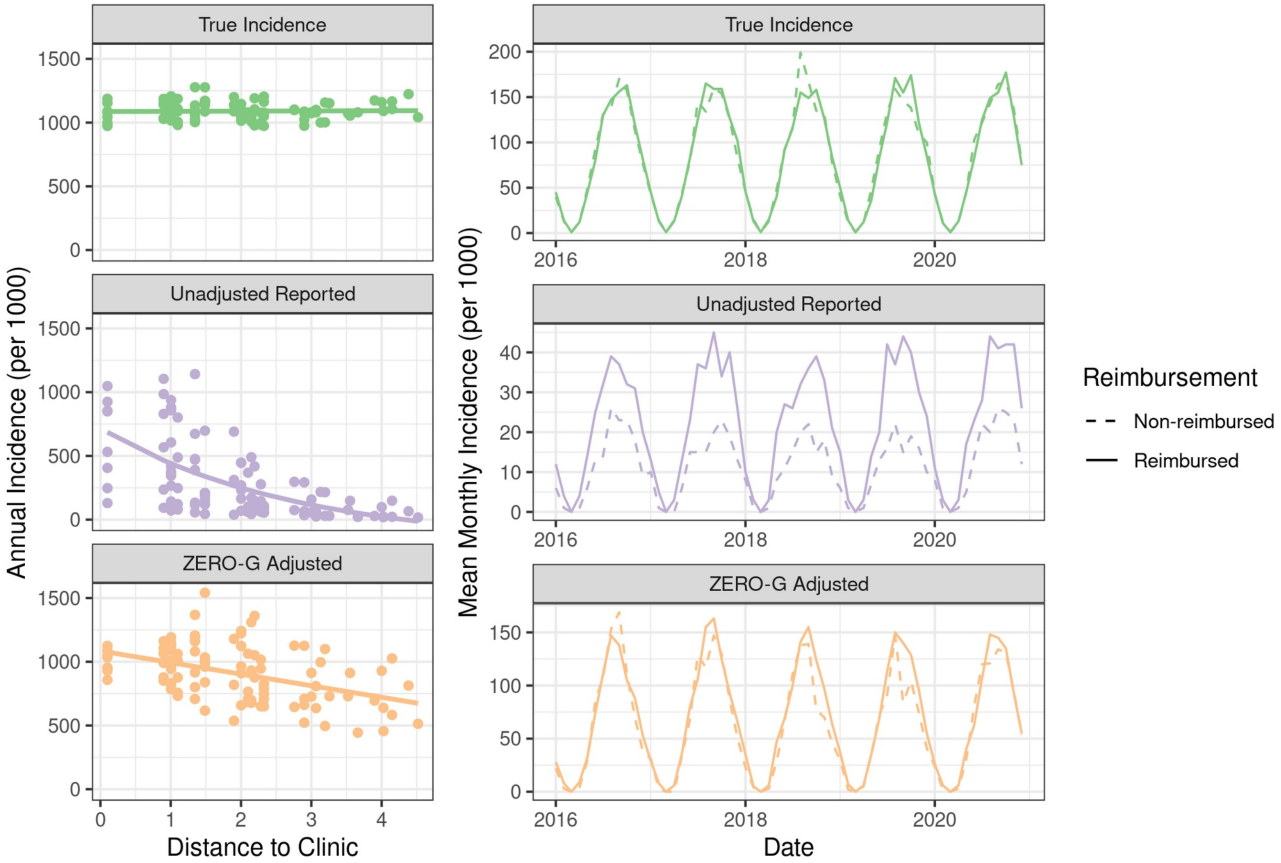


**Figure S1.7.** Biases due to geographic location and financial policies were reduced in the ZERO-G adjusted data relative to the unadjusted data. Left: The average annual incidence per patch relative to a zone's distance to the nearest clinic. Right: The mean monthly incidence in zones with fee reimbursement and zones without fee reimbursement policies.

### 2. Case Study: Malaria Incidence in Ifanadiana, Madagascar

**Table S2.1.** Table of best fit parameters estimated via MLE in the estimation of healthcare access A via Equations 7- 12.

| | | Age Class | | |
|---|---|---|---|---|
| **Parameter** | **Description** | **Children** | **Juvenile** | **Adult** |
| $\beta_{s1}$ | PHC coefficient: initial Pivot intervention | 9.360 | 9.124 | 25.248 |
| $\beta_{s2}$ | PHC coefficient: fee reimbursement | 17.274 | 19.163 | 34.732 |
| $\beta_{s3}$ | PHC coefficient: PHC type | 0.049 | 0.099 | -0.285 |
| $\beta_{s4}$ | PHC coefficient: Number of staff | 2.069 | 1.386 | 2.584 |
| $\beta_{s5}$ | PHC coefficient: Distance to District | 6.475 | 2.833 | 3.743 |
| $\lambda$ | Distance decay | 0.117 | 0.109 | 0.096 |
| $\beta_{\lambda}$ | Distance decay shape parameter | -0.278 | -0.075 | -0.26 |
| $\beta_C$ | Competition for PHC services coefficient | 0.090 | 0.068 | 0.058 |
| $\beta_{g1}$ | Time coefficient: linear trend | -0.003 | -0.003 | 0.002 |
| $\beta_{g2}$ | Time coefficient: seasonal trend | 0.369 | -0.572 | 0.168 |
| $\phi$ | Time coefficient: month offset | 12.801 | 7.477 | -9.633 |

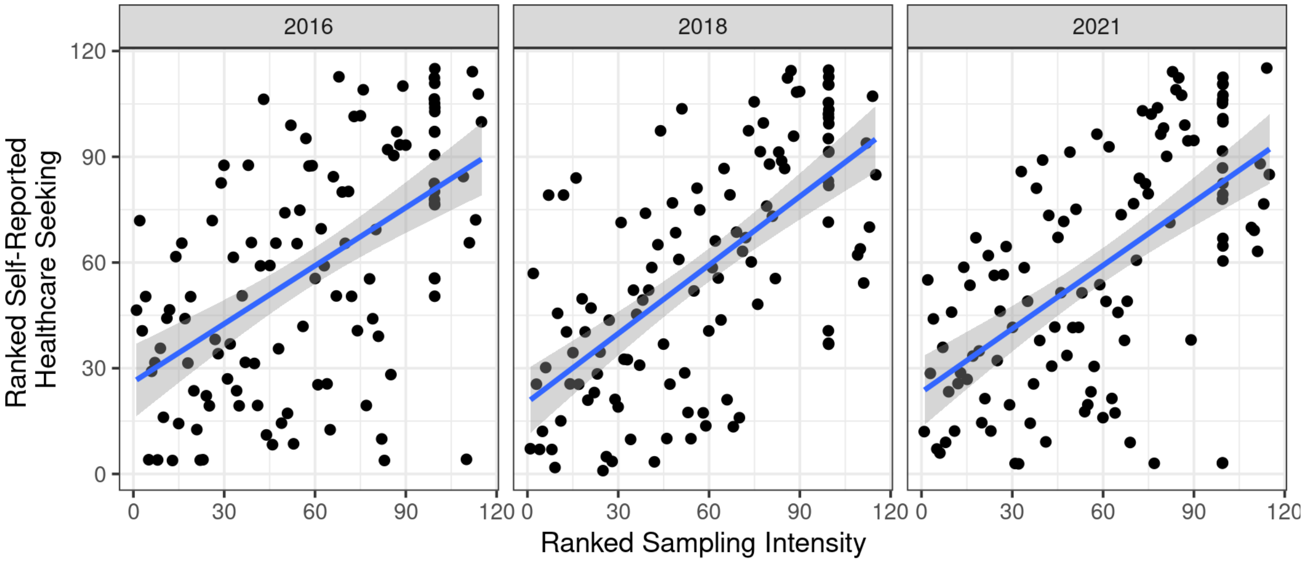


**Figure S2.1** Scatter plots illustrating the relationship between sampling intensity estimated via the floating catchment area (FCA) method and the self-reported healthcare seeking rates from the IHOPE cohort for three survey years (2016,2018,2021).
